# Neurobiology-based Cognitive Biotypes Using Multi-scale Intrinsic Connectivity Networks in Psychotic Disorders

**DOI:** 10.1101/2024.05.14.24307341

**Authors:** Pablo Andrés-Camazón, Covadonga Martínez Diaz-Caneja, Ram Ballem, Jiayu Chen, Vince D. Calhoun, Armin Iraji

## Abstract

**Objective:** Understanding the neurobiology of cognitive dysfunction in psychotic disorders remains elusive, as does developing effective interventions. Limited knowledge about the biological heterogeneity of cognitive dysfunction hinders progress. This study aimed to identify subgroups of patients with psychosis with distinct patterns of functional brain alterations related to cognition (cognitive biotypes).

**Methods:** B-SNIP consortium data (2,270 participants including participants with psychotic disorders, relatives, and controls) was analyzed. Researchers used reference-informed independent component analysis and the NeuroMark 100k multi-scale intrinsic connectivity networks (ICN) template to obtain subject-specific ICNs and whole-brain functional network connectivity (FNC). FNC features associated with cognitive performance were identified through multivariate joint analysis. K-means clustering identified subgroups of patients based on these features in a discovery set. Subgroups were further evaluated in a replication set and in relatives.

**Results:** Two biotypes with different functional brain alteration patterns were identified. Biotype 1 exhibited brain-wide alterations, involving hypoconnectivity in cerebellar-subcortical and somatomotor-visual networks and worse cognitive performance. Biotype 2 exhibited hyperconnectivity in somatomotor-subcortical networks and hypoconnectivity in somatomotor-high cognitive processing networks, and better preserved cognitive performance. Demographic, clinical, cognitive, and FNC characteristics of biotypes were consistent in discovery and replication sets, and in relatives. 70.12% of relatives belonged to the same biotype as their affected family members.

**Conclusions:** These findings suggest two distinctive psychosis-related cognitive biotypes with differing functional brain patterns shared with their relatives. Patient stratification based on these biotypes instead of traditional diagnosis may help to optimize future research and clinical trials addressing cognitive dysfunction in psychotic disorders.

## INTRODUCTION

Cognitive dysfunction is widely recognized as a transdiagnostic core dimension of psychotic disorders and is consistently associated with general functional outcomes. The effectiveness of current therapeutic or preventive strategies for cognitive dysfunction is limited (2,3) and the development of new treatments is hampered by our scarce knowledge about the biological underpinnings of cognitive dysfunction (4). Distinct patient subgroups with different neurobiology (i.e., biotypes) may exist, necessitating different treatments (4–6). Nevertheless, DSM diagnoses are based on different patterns of psychotic or affective symptoms, which do not necessarily reflect the brain alterations that may underlie cognitive dysfunction in these disorders. Cognitive dysfunction-associated brain alterations could be different in people with the same DSM diagnosis or overlap between people with a different diagnosis or with their first-degree relatives, known to present intermediate cognitive performance (7). Despite this, cognitive interventions are usually developed for participants with a specific DSM diagnosis (8), rather than for biotypes based on neurobiology. This method ignores the potential biological heterogeneity underlying cognitive dysfunction and poses a substantial impediment to its success (4,5).

Previous works in this area have been limited mainly to three categories. The first category has focused on identifying subgroups using observable cognitive performance (9). This methodology resembles DSM, through computational approaches, but probably with the same underlying caveats. The second category has identified general subgroups combining cognitive performance and laboratory tasks (10,11). These categories often fail to leverage the neural underpinnings of cognition. The third category has utilized magnetic resonance imaging (MRI) to identify subgroups based on brain characteristics, but their emphasis has been on identifying general subgroups rather than oriented toward cognitive dysfunction (12–17). Covariation patterns between brain features and symptom dimensions may vary, indicating distinct brain mechanisms for each dimension. Relying solely on general subgroups may obscure again the underlying biological diversity of cognitive dysfunction, limiting the utility of these subgroups in the understanding of cognitive dysfunction.

Recent research has shown that resting-state functional connectivity (rsfMRI) (18) exhibits stronger associations with cognitive performance than structural MRI (19). Consequently, rsfMRI emerges as a more suitable tool for the discovery of cognition-related neurobiology-based biotypes of psychotic disorders.

Recent advancements have led to a standardized and fully automated framework, called Neuromark (20) for identifying functional patterns across participants and datasets in rsfMRI. This includes a canonical template with 105 multi-scale intrinsic connectivity networks (ICNs) from data collected from over 100,000 participants (21). This template can be used along with reference-informed independent component analysis (ICA) (22,23) to identify these ICNs in new participants and to compute their functional network connectivity (FNC).

In this work, we hypothesize that subgroups of patients or biotypes with psychotic disorders with different patterns of FNC related to cognitive dysfunction and different cognitive profiles can be identified with this method. We use a large dataset of participants with schizophrenia (SZ), schizoaffective disorder (SAD), and bipolar disorder with psychotic symptoms (BDP), their first-degree relatives, and controls. We will validate the subgroups in a replication sample and in first-degree relatives.

## METHODS

### 1. Participants

We analyzed data from 2270 participants recruited by the Bipolar-Schizophrenia Network on Intermediate Phenotypes (B-SNIP) Consortium 1 and 2. Patients meeting the criteria for BDP, SAD, or SZ were included (N=1179), and their first-degree relatives (N=465). Controls (N=626) had no history of lifetime psychosis syndromes, recurrent mood syndromes, or first-degree relatives with psychosis. We used all available data from participants who had completed rsfMRI and Brief Assessment of Cognition in Schizophrenia (BACS) data **(Table S1)**.

### 2. Discovery and replication sets

Our sample was divided into discovery and replication sets comprising 80% and 20% of the sample. The proportion of patients, relatives, and controls was consistent in both sets.

### 3. Clinical and cognitive assessments

Participants meeting a diagnosis for SZ, SAD, or BDP were rated on clinical scales. All participants were rated on the BACS. We also incorporated the Weschler Memory Scale Backward and Forwards subtests for the cognitive assessment (24). For patients recruited in B-SNIP2, information regarding childhood learning difficulties was collected.

### 4. Estimating subject-specific multi-scale intrinsic connectivity networks (ICNs) and functional network connectivity (FNC)

Imaging data acquisition and preprocessing details can be found in supplementary material. We used the GIFT software toolbox (http://trendscenter.org/software/gift) (25) to perform multivariate-objective optimization ICA with reference (MOO-ICAR) (21,22,26), and generate subject-specific ICNs. As a reference, we used the Neuromark_fMRI_2.0 template (http://trendscenter.org/data), which includes highly replicated 105 ICNs across different spatial scales (21) in over 100k individuals. Next, we computed subject-level static FNC by calculating pairwise Pearson correlations between the cleaned time courses of ICNs. This process resulted in a 105 × 105 symmetric FNC matrix for each participant, which represents the whole-brain functional connectome (27,28) **(Fig. 1A)**.

**Fig. 1.**
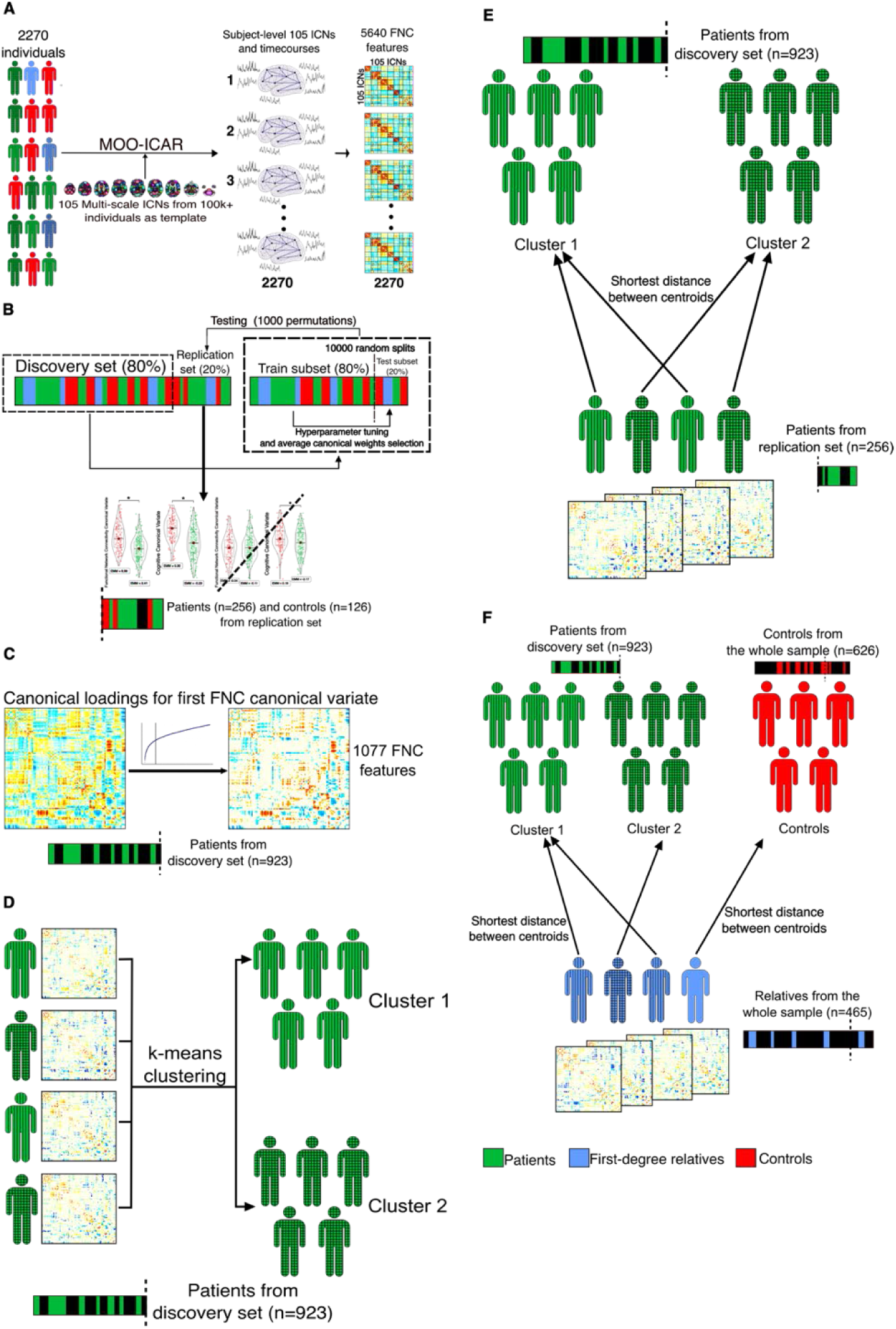
Overview of the analytic pipeline. **A)** We employed multivariate-objective optimization independent component analysis with reference (MOO-ICAR) to estimate 105 multi-scale intrinsic connectivity networks (ICNs) at the subject level. The reference for the 105 ICNs was derived from a large sample of over 100,000 participants. We computed subject-level static functional network connectivity (FNC) by calculating pairwise Pearson correlations between cleaned time courses of ICNs, resulting in a 105 × 105 symmetric FNC matrix for each participant. ICNs are grouped based on their anatomical and functional properties. **B)** Principal Component Analyses Plus Canonical Correlation (PCA-CCA) was fitted on the discovery set, including patients, relatives, and controls, to find FNC features associated with the Brief Assessment of Cognition in Schizophrenia and Wechsler Memory Scale Backward and Forwards Tests. The optimal number of principal components was estimated directly from the data Elbow criteria. Three canonical correlations remained statistically significant in the replication set and two remained statistically significant after adjusting for covariates. We conducted a pair-wise comparison between patients and controls for the remaining two canonical variates adjusting for covariates in the replication set. Patients and controls presented statistically significant differences in both cognitive canonical variates and in the first FNC canonical variate but did not in the second FNC canonical variate, therefore we did not include the second pair in subsequent analyses. **C)** We selected FNC features with the highest correlation with the first canonical variate in patients in the discovery set. We selected FNC features with a loading >|0.1047| according to the elbow method *(Cognitive FNC features in Psychosis* or CFPs). Left: Correlation values of the 5460 functional network connectivity features and the first FNC canonical variate in 105 × 105 symmetric matrices in patients in the discovery set. Blue colors: negative correlation. Yellow-red colors: positive correlation. Lighter colors: correlation closer to 0. Middle: elbow plot of ranked absolute values of correlations with a vertical line at 0.1047, the value chosen as threshold. Right: FNC features with a correlation with absolute values lower than 0.1047 and therefore not included in k-means clustering are shown in white. The 1077 FNC features selected for k-means clustering are shown in color. **D)** We conducted k-means clustering using patients from the discovery set to find subgroups of patients based on CFPs. Silhouette index solution for two clusters was statistically significant (p=0.0005). **E)** We assigned patients from the replication set to one of the clusters obtained in the discovery set based on the shortest Euclidean distance between each subject’s centroid and the clusters’ centroid for the 1077 CFPs. **F)** We computed the centroid of the 1077 CFPs in the control group to simulate a control cluster. Like E, we assigned first-degree relatives to either one of the patients’ clusters or to the control cluster based on the shortest distance between the centroid of each subject and the centroids of the clusters.

### 5. Canonical FNC signatures of cognitive performance

First, we aimed to find FNC features highly associated with cognitive performance (BACS and Weschler Memory Scale Backward and Forwards tests). Secondly, we used these features to identify cognition-related neurobiology-based subgroups of patients or biotypes. For this purpose, we used a Principal Component Analysis Plus Canonical Correlation Analyses (PCA-CCA) model. PCA-CCA finds pairs of cognitive and FNC weights (canonical weights) such that the linear combination of the cognitive and FNC variables maximizes the correlation (canonical correlation) between the resulting pair (canonical pair) of latent variables (canonical variates). PCA-CCA was performed on the discovery set, including patients, relatives, and controls **(Fig. 1B)**. By combining the three groups we achieved a larger sample size, boosting the statistical power and mitigating overfitting (19,32). This is also crucial as it allows us to select the canonical variates with significant differences between participants with psychosis and controls. By focusing on these features, subsequent clustering analysis has a higher likelihood of identifying subgroups of patients based on FNC features related to cognitive dysfunction in psychosis, rather than simply capturing general population variability. The optimal number of principal components was estimated directly from the data using the Elbow method ensuring that we retained relevant information while achieving a well-posed and more generalizable model. We included further methods to optimize the model and improve the robustness of our findings (supplementary material, **Fig. 1B**). We tested canonical correlations’ statistical significance in replication and discovery sets with permutation analyses (10000 permutations).

Next, we examined the influence of covariates (age, sex, race, socioeconomic status, and chlorpromazine equivalents) in the association between canonical variates in the replication set. For this, we conducted linear models with canonical cognitive variates (CV_Cog_) as dependent variables and canonical FNC variates (CV_FNC_) and covariates as independent variables. Canonical pairs without statistically significant associations in the replication set in these linear models were excluded from subsequent analyses.

### 6. Determining cognition-related FNC features in participants with psychosis

Next, we identified cognition-related FNC features in psychosis. First, we applied group comparison between patients and controls while controlling for the same previous covariates to identify canonical pairs with significant differences between groups in both canonical variates in the replication set (which we called psychosis-related) **(Fig. 1B)**. Since we were aware of possible problems associated with covariates correlated with one group (33) we repeated these analyses excluding chlorpromazine equivalents (associated with participants with psychosis) and obtained similar results. Subsequently, we calculated Pearson correlations between this CV_FNC_ and FNC features in the patient’s group. By this, we pinpointed FNC features that exhibit covarying patterns similar to the psychosis-related CV_FNC_ across participants with psychotic disorders. The correlation values were ranked, and the elbow method was used to establish a threshold. We selected the FNC features with higher correlations than |0.1047| **(Fig. 1C)**. We refer to these FNC features as Cognitive FNC Features in Psychosis or CFPs **(Fig. 1C)**.

### 7. Identifying cognition-related psychosis biotypes

We evaluated whether CFPs conformed to a distribution with potential underlying clusters in the patient’s group in the discovery set. We applied k-means clustering with Euclidean distance **(Fig. 1D)**. We computed the silhouette index to find the optimal number of clusters (k) and its statistical significance against the null hypothesis of a distribution with no clusters (32). Clustering stability was then evaluated using a bootstrapping resampling technique (*n* bootstraps=1000) and Jaccard similarity average (34). We further considered these clusters as biotypes.

### 8. Biotypes validation and characterization

In k-means clustering, each cluster is represented by its centroid. This enables the assignment of a new subject to one of the established clusters based on the similarity between the CFPs of the subject and the cluster centroids (shortest Euclidean distance between subject and cluster centroids) (35,36). We assigned patients from the replication set to clusters obtained in the discovery set using this method (**Fig. 1E**).

First-degree relatives present intermediate cognitive performance between patients and controls (34, **Table S1**), therefore, we hypothesized that some relatives present more similar CFP patterns to their affected family member than to controls, while other relatives may be more similar to controls than to patients. To evaluate this, we computed the centroid representative for the control group (control cluster or biotype) by calculating the average value of their CFPs. Relatives were then assigned to either one of the patients’ clusters or the control cluster based on their closest centroid (**Fig. 1F**). We calculated the proportion of relatives assigned to the same cluster as their affected family members. We compared clusters/biotypes of patients from discovery and replication sets and clusters/biotypes of relatives, for demographic, clinical, cognitive variables, and FNC features. We excluded first-degree relatives with a psychotic disorder.

### 9. Comparison with DSM diagnostic categories

Using a consistent approach, we computed the centroids for SZ and BPD participants, assigning their relatives to one of these diagnostic groups or control group. We hypothesize that DSM diagnoses exhibit heterogeneity and overlap in FNC features related to cognition, therefore, the proportion of relatives in the same cluster will be lower than when using biotypes.

For detailed methodological and complementary information see the corresponding sections in the supplementary material.

## RESULTS

Demographic, clinical, and cognitive characteristics of discovery and replication sets are shown in **Table S1.**

### 1. Canonical FNC signatures of cognitive performance

Three canonical pairs presented statistically significant correlations in both the discovery (r_Dis1_=0.49, p_Dis1_=0.001; r_Dis2_=0.34, p_Dis2_<0.001; r_Dis3_=0.33, p_Dis3_<0.001, **Fig. 2A**) and replication set (r_Rep1_=0.47, p_Rep1_=0.001; r_Rep2_=0.24, p_Rep2_<0.001; r_Rep3_=0.13, p_Rep3_=0.002, **Fig. 2B**). Loadings are shown in **Fig. 2C and 2D**. Associations in the first and second pairs remained statistically significant after adjusting for covariates in the replication set (β_Rep1_=0.24, CI_Rep1_: 0.16 – 0.33, p_Rep1_<0.001; β_Rep2_=0.12, CI_Rep2_: 0.02 – 0.21, p_Rep2_=0.015), but third pair association did not (β_Rep3_=0.03, CI_Rep3_: −0.07 – 0.14, p_Rep3_=0.500). The third pair was not included in subsequent analyses (see supplementary material).

**Fig. 2.**
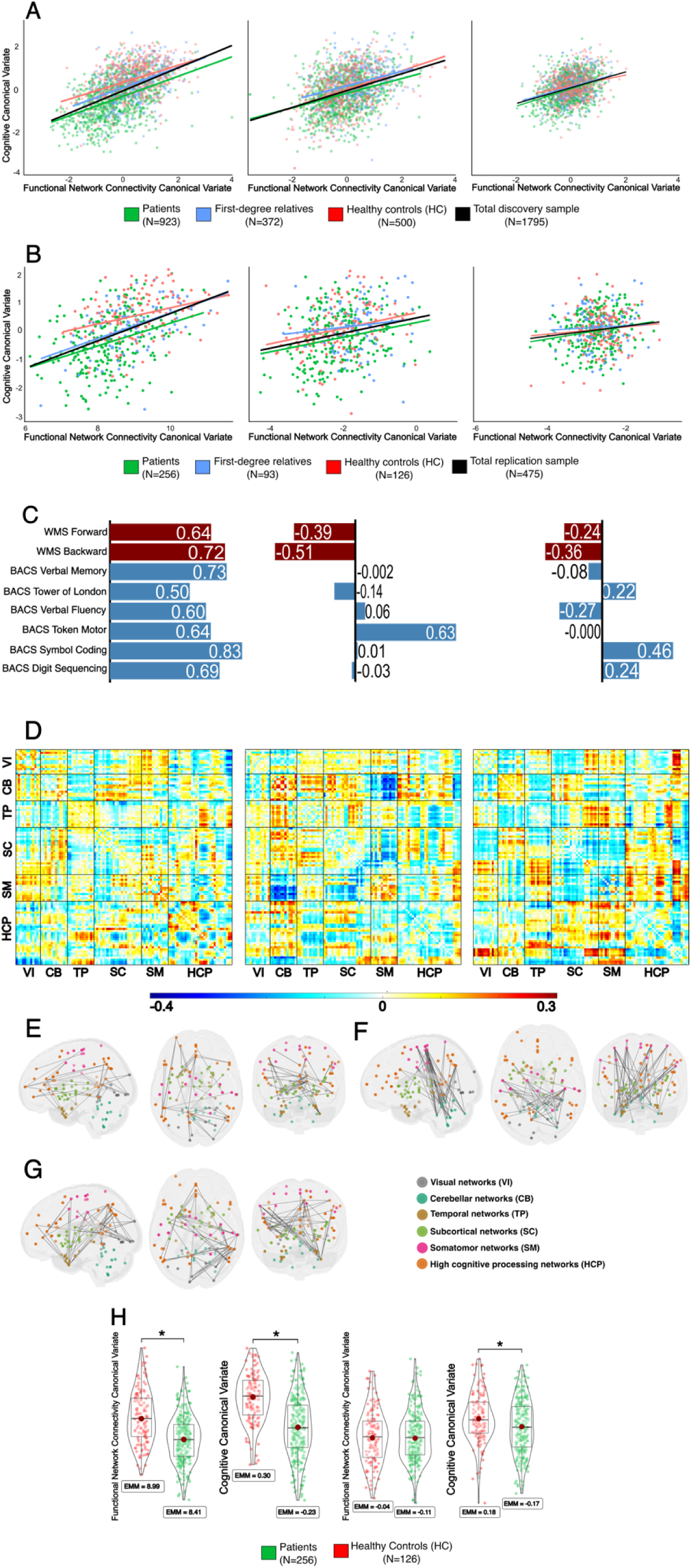
Canonical correlations and canonical loadings between cognitive performance and functional network connectivity features. **A)** Scatterplots of the first (left), second (center), and third (right) canonical pairs in the discovery set. Canonical correlations: r_Dis1_=0.49, p_Dis1_=0.001; r_Dis2_=0.34, p_Dis2_=0.001; r_Dis3_=0.33, p_Dis3_=0.001. P-values computed with 1000 permutations **B)** Scatterplots of the first (left), second (center), and third (right) canonical pairs in the replication set. Canonical correlations: r_Rep1_=0.47, p_Rep1_<0.001; r_Rep2_=0.24, p_Rep2_<0.001; r_Rep3_=0.13, p_Rep3_=0.002. **C)** Loadings (Pearson correlation between cognitive subtests and cognitive canonical variates) for Brief Assessment of Cognition in Schizophrenia (BACS, blue) subtests and Weschler Memory Scales (WMS, red) Backward and Forward for the first (left), second (center) and third (right) canonical pairs. **D)** Loadings (Pearson correlation between FNC and FNC canonical variates) for the 5460 FNC and the first (left), second (center), and third (right) canonical pairs. Loadings are represented in 105×105 symmetric matrices. The 105 multiscale intrinsic connectivity networks are grouped as follows: Visual networks (VI), cerebellar networks (CB), temporal networks (TP), subcortical networks (SC), somatomotor networks (SM), and high cognitive processing networks (HCP). **E, F and G)** Brain maps in MNI152 space, where nodes are the coordinates of peak activation points of the 105 intrinsic connectivity networks and edges are 10% functional connectivity features (FNC) with the highest loadings for each FNC canonical variate (E, first; F, second; G, third**). H)** Violin plots of participants with psychosis and controls pairwise comparisons of the first (left) and second (right) canonical pairs in the replication set. Two-tailed t-tests obtained from linear models adjusting for covariates: First functional network canonical variate, d=0.58, t(466)=5.28, p_adj_<0.0001; First cognitive canonical variate, d=0.53, t(466)=5.05, p_adj_<0.0001; Second functional network connectivity canonical variate, d=0.07, t(466)=0.56, p_adj_=0.84; Second cognitive canonical variate, d=0.35, t(466)=3.05, p_adj_=0.007. t: t-statistic, t(degrees of freedom), d: difference. * statistically significant differences between groups. EMM: Estimated Marginal Mean. P-value: Tukey method for comparing a family of 3 estimates.

### 2. Determining cognition-related FNC features in participants with psychosis

Pairwise comparison in the replication set (**Fig. 2H**) showed statistically significant differences between patients and controls in CV_FNC1_ (controls–patients, d=0.58, t(466)=5.28, p_adj_<0.0001), CV_Cog1_ (controls–patients, d=0.53, t(466)=5.05, p_adj_<0.0001), and CV_Cog2_ (controls–patients, d=0.35, t(466)=3.05, p_adj=_0.007), but not in CV_FNC2_ (controls–patients, d=0.07, t(466)=0.56, p_adj_=0.84). Therefore, we included the first pair but not the second in subsequent analyses since we were interested in canonical pairs with differences between patients and controls in both cognition and FNC. We selected FNC features with the highest correlation with the CV_FNC1_ in patients in the discovery set (CFPs) for k-means clustering. Elbow’s method suggested 0.1047 as a threshold, comprising 1077 FNC features **(Fig. 1C)**.

### 3. Identifying cognition-related psychosis biotypes, validation, and characterization

We ran k-means clustering to identify two clusters **(Fig. 1D)**. Silhouette index (0.05) solution for two clusters was statistically significant (p=0.0005). Jaccard similarity values were 0.935 for Cluster 1 and 0.921 for Cluster 2. A value higher than 0.85 suggests a highly stable cluster (29, 31). Of those first-degree relatives with their family members included in analyses, 76.56% (N=281/367) were assigned to one of the patient’s biotypes, and of those relatives, 70.12% (N=197/281, χ2=7.03, p<0.00001) were assigned to the same biotype as their family member **(Fig. 1F)**. 23.4% (N=86/367) of relatives were assigned to the control biotype (see supplementary material). In the same analyses with BDP and SZ diagnosis, only 54.85% of relatives assigned to psychosis clusters were assigned to the same diagnosis cluster as their family members (N=96/175, χ2=1.22, p=0.269).

#### 3.1 Patient biotypes in the discovery set

Biotype 1 exhibited more extensive FNC alterations than biotype 2. Biotype 1 displayed hypoconnectivity between visual-somatomotor, cerebellar-subcortical, high cognitive processing-cerebellar, somatomotor-subcortical, and high cognitive processing-temporal networks, and hyperconnectivity between visual-subcortical, visual-cerebellar, somatomotor-subcortical and high cognitive-somatomotor networks (**Fig.3A and B)**. Biotype 2 displays hypoconnectivity between high cognitive processing-somatomotor, high cognitive processing-subcortical and subcortical-cerebellar, and hyperconnectivity between cerebellar-somatomotor, subcortical-somatomotor, and temporal-high cognitive processing networks (**Fig.3A and B)**. Compared to biotype 2 and after false discovery rate correction (FDR), biotype 1 presented a statistically significantly higher proportion of African Americans and lower of Caucasians, a higher proportion of SZ and lower of BDP, older age, lower socioeconomic status, more similarity to SZ than BDP (schizo-bipolar scale, SBS), a lower but small difference in global and social functioning (GAF and BSFS), worse cognitive performance across all cognitive subtests and a higher proportion of patients with a history of childhood learning difficulties in all categories **(Fig.4**, **Fig.5A and Table S5)**.

**Fig. 3.**
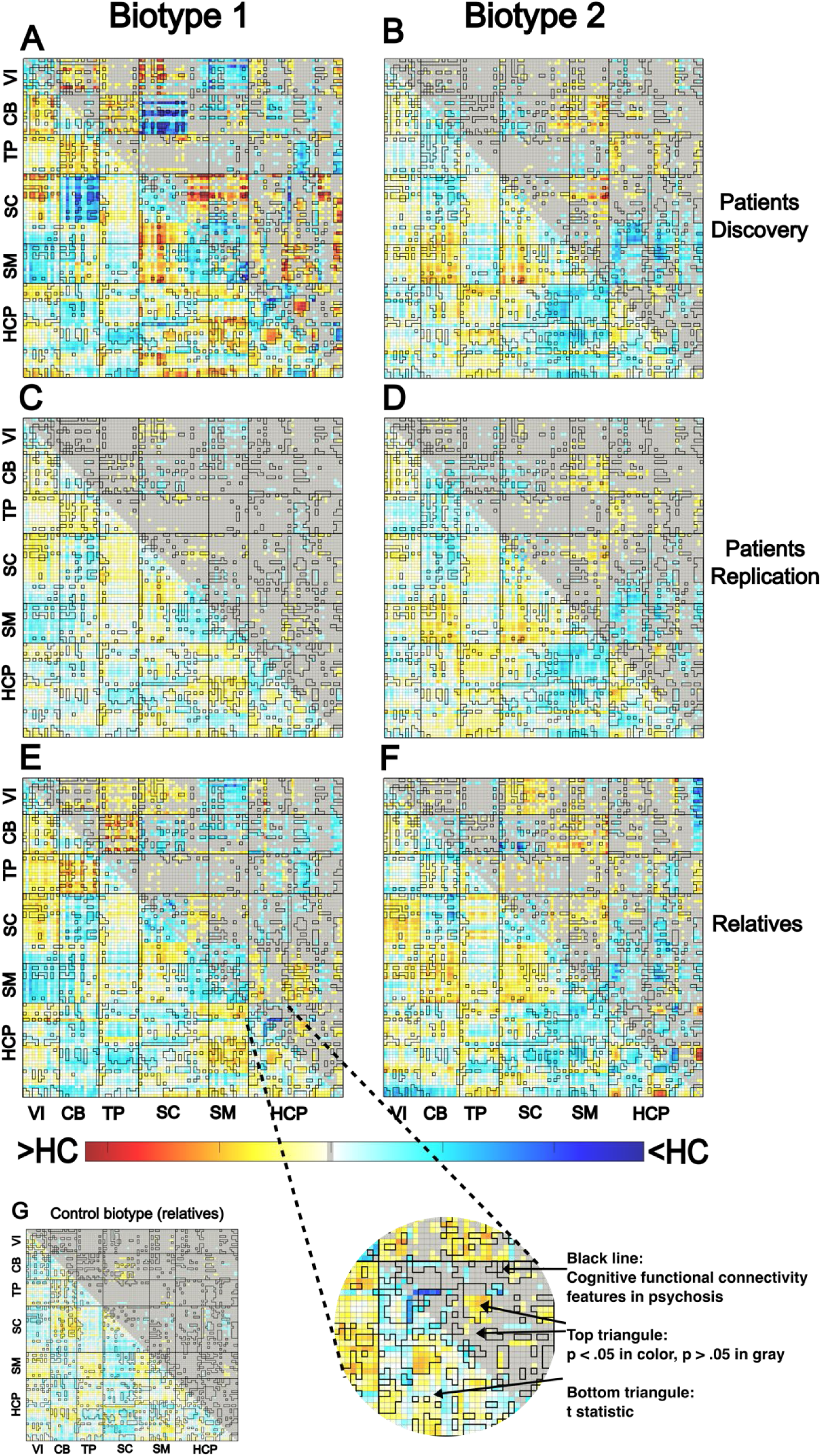
Matrices for the differences in functional network connectivity between controls (N = 626) and biotypes. Discovery set: biotype 1, N=426; biotype 2, N=497. Replication set: biotype 1, N=110, biotype 2 N=146. First-degree relatives: biotype 1, N=153; biotype 2, N=178, cognitive biotype, N=95. For each functional network connectivity (FNC) feature, we fit a linear model adjusting for sex, age, race, ethnicity, site, and head motion and conducted a two-tailed t-test to compare controls with each biotype of patients and relatives. Bottom-left triangle: t-statistic. Top-right triangle: p-values, those that reached statistical significance (p<0.05) after false discovery rate (FDR) correction are shown with colors (–log10(p-value)×sign(t-statistic) scale); otherwise, they are shown in gray. Yellow-Red colors: higher FNC in patients/relatives compared to controls. Blue colors: lower FNC in patients/relatives compared to controls. *Cognitive FNC features in psychosis (*CFPs) included in k-means clustering are shown with a black line. Visual networks (VI), cerebellar networks (CB), temporal networks (TP), subcortical networks (SC), somatomotor networks (SM), and high cognitive processing networks (HCP). First-degree relatives with a diagnosis of a psychotic disorder were excluded from these analyses.

**Fig. 4.**
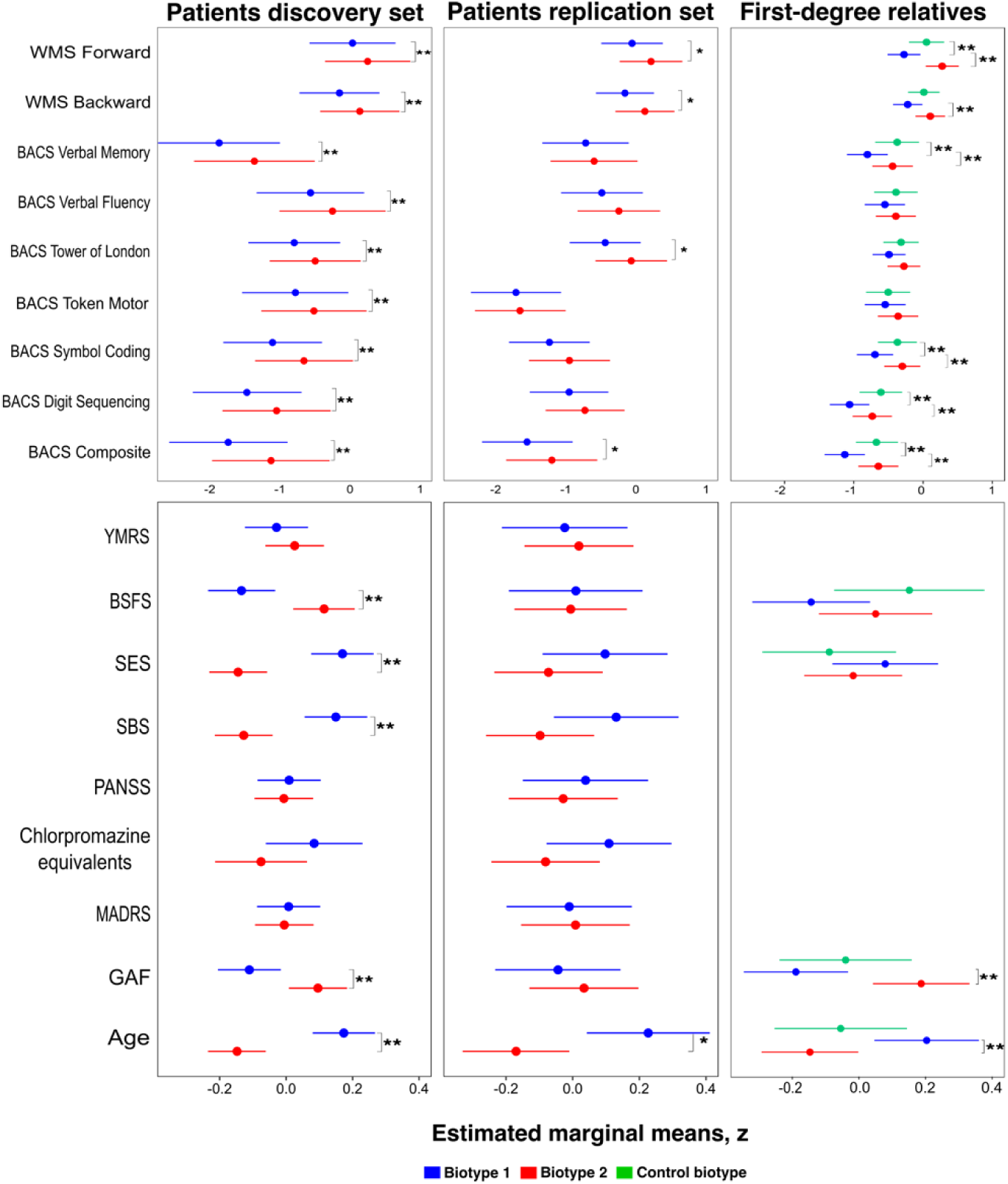
Comparison of demographic, clinical and cognitive characteristics of biotypes (I). Discovery set: biotype 1, N=426; biotype 2, N=497. Replication set: biotype 1, N=110, N=146. First-degree relatives: biotype 1, N=153; biotype 2, N=178, control biotype, N=95. WMS: Weschler Memory Scale; BACS: Brief Assessment of Cognition in Schizophrenia; YMRS: Young Mania Rating Scale; BSFS: Birchwood Social Functioning Scale; SES: Socioeconomic Status (Hollingshead index); SBS: Schizo-bipolar Scale; PANSS: Positive and Negative Syndrome Scale for Schizophrenia; Chlorpromazine equivalents: Average daily chlorpromazine dose; MADRS: Montgomery-Asberg Depression Rating Scale; BACS: Brief Assessment of Cognition in Schizophrenia; BSFS: Birchwood Social Functioning Scale; SES: Socioeconomic status; SBS: Schizo-bipolar Scale; Chlorpromazine equivalents: Average daily chlorpromazine dose; MADRS: Montgomery-Asberg Depression Rating Scale; GAF: Global Assessment of Functioning Scale; Z-scores are shown. BACS z-scores were obtained from normative data stratified by age and sex. * Non-adjusted p-value < 0.05 from a two-tailed t-test; ** False discovery rate correction for multiple testing p-value <0.05. Comparisons with three biotypes in first-degree relatives were adjusted with the Tukey method for comparing a family of three estimates. WMS: Statistics were obtained from linear models that also accounted for the influence of age, sex, race, ethnicity, site, and socioeconomic status. BACS: Statistics were obtained from linear models that also accounted for the influence of race, ethnicity, site, and socioeconomic status (age and sex accounted for when computing z-scores). Statistical details are shown in Table S5. First-degree relatives with a diagnosis of a psychotic disorder were excluded from these analyses.

**Fig. 5.**
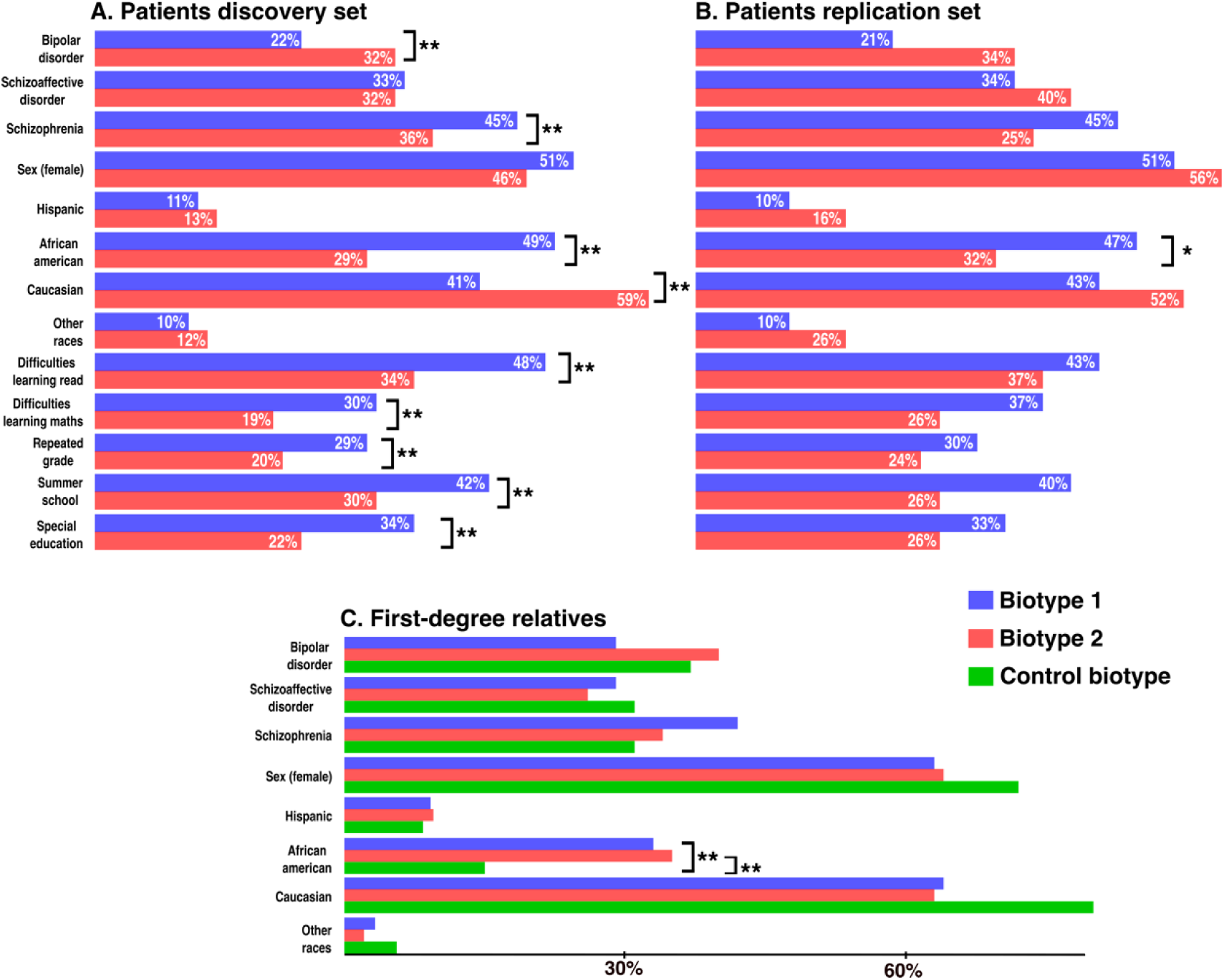
Comparison of demographic and clinical characteristics (II) and childhood learning difficulties between biotypes. Discovery set: biotype 1, N=426; biotype 2, N=497. Replication set: biotype 1, N=110, N=146. First-degree relatives: biotype 1, N=153; biotype 2, N=178, control biotype, N=95. * Non-adjusted p-value < 0.05 from Pearson’s Chi-squared test; ** False discovery rate correction for multiple testing p-value <0.05. DSM diagnosis in first-degree relatives refers to the diagnosis of the affected family member. Statistical details are shown in Table S5. First-degree relatives with a diagnosis of a psychotic disorder were excluded from these analyses.

#### 3.2 Patient biotypes in the replication set

Fewer differences in FNC, demographic, clinical, and cognitive characteristics reached statistical significance, but a similar pattern of differences in the same direction was observed **(Fig.3C and D, Fig.4 and Fig.5B).** African Americans and older age were statistically significant. Higher similarity to SCZ (SBS) was close to significance (p=0.07) **(Fig.4 and Table S5)**. Regarding cognition, similar to the discovery set, biotype 1 displayed an average worse performance in all cognitive subtests and a higher proportion of patients with learning difficulties during childhood in all categories, but only differences in BACS composite, tower of London, WMS backward, and forward tests reached statistical significance. Symbol coding was close to significance (p=0.054). None survived FDR correction **(Fig. 4**, **Fig.5B, and Table S5)**.

#### 3.3 First-degree relatives biotypes

Congruent patterns of differences also emerged in first-degree relatives **(Fig.3E, F and G, Fig. 4**, **Fig.5C, and Table S5)** in FNC, demographic and clinical characteristics. Biotype 2 also showed hypoconnectivity between visual-high cognitive performance networks and hyperconnectivity between visual-subcortical networks, a pattern not observed in the same biotype in patients. As expected, the control biotype presented substantially fewer FNC differences. After FDR correction and compared to biotype 2, biotype 1 exhibited statistically significant older age, lower GAF, and a higher proportion of African Americans. Biotype 1 also presented worse cognitive performance across all subtests. Differences in the BACS composite, verbal learning, digit sequencing, symbol coding, and WMS backward and forward subtests were statistically significant and survived FDR. Compared to relatives in the control biotype, biotype 1 displayed worse performance in BACS composite, verbal memory, symbol coding, digit sequencing, and WMS forwards subtests after FDR. Biotype 2 (more cognitively preserved in patients) and control biotype did not exhibit statistically significant differences in cognitive performance. This pattern mirrors the findings observed in patients.

## DISCUSSION

Our study aimed to identify neurobiology-based cognition-related biotypes in patients with psychotic disorders, characterized by distinct functional brain alterations related to cognition. Firstly, we identified a robust multivariate correlation between brain-wide FNC and cognitive performance across patients, relatives, and controls in a never-seen replication set. We found a higher correlation than previously reported (19), potentially attributable to methodological improvements through our constrained ICA NeuroMark approach and the multi-scale ICNs template. Patients exhibited significantly lower scores in the first FNC and cognitive canonical variates and remained significant after adjusting for covariates such as antipsychotic medication. This is in line with previous studies that suggest that disruptions in brain networks are implicated in cognitive dysfunction in psychosis (39,40).

Secondly, we identified two biotypes with distinct FNC characteristics linked to cognitive performance (CFPs). Biotype 1 consistently exhibited poorer cognitive performance and a higher prevalence of childhood learning difficulties in both discovery and replication sets. Lack of statistically significant differences in cognitive performance and other characteristics between biotypes in the replication set is likely due to limited statistical power. This is supported by similar patterns in demographic, clinical, cognitive, and FNC characteristics between biotypes in discovery and replication sets. Analogous patterns were observed in first-degree relatives: biotype 1 displayed inferior cognitive performance in six cognitive subtests compared to biotype 2 and in five compared to the control biotype (first-degree relatives more similar to controls), with similar patterns in FNC differences compared to controls.

The two identified biotypes potentially represent distinct subgroups of patients with divergent patterns of brain networks implicated in cognitive dysfunction and spanning widely across the brain. This different pattern of hypo-hyper connectivity (reversed in somatomotor-high cognitive processes, temporal-high cognitive processes, and subcortical-high cognitive processes networks to a different extent) may suggest that biotypes are not only different in severity but also in brain alterations or compensation mechanisms. The involvement of visual networks is noteworthy, as their implication in psychosis is debated (41).

To our knowledge, this study represents the first attempt to identify neurobiology-based psychosis biotypes associated with a specific symptom dimension in both patients and relatives. We consider this approach as a potential improvement for unraveling the biological heterogeneity within psychosis compared to previous efforts. The identification of two patient biotypes —one with more pronounced brain alterations and poorer cognitive performance— aligns with findings from other studies (15,16) that utilized different methodology and structural MRI. The convergence of findings potentially suggests the validity for both approaches.

Our third key finding reveals that 70.12% (χ2=7.0297, p<0.00001) of relatives were assigned to the same biotype as their family members with psychosis, suggesting that relatives present similarities in the functional brain patterns related to cognition that delineate biotypes. Shared genetic and environmental backgrounds likely contribute to this phenomenon. Consistent differences in childhood learning difficulties prevalence between the discovery and replication sets (though reaching statistical significance only after FDR in the discovery set), suggest potential biotype-related distinctions in cognitive performance present, at least, since childhood. This is in line with the neurodevelopmental hypothesis of psychosis. Notably, biotype 1 showed a higher representation of African Americans among patients and relatives, aligning with research on childhood adversity disparities in African Americans impacting structural brain differences (42). This overrepresentation may reflect adverse environments for biotype 1, potentially influencing cognitive development, leading to learning difficulties since childhood and more pronounced FNC disruptions in adulthood.

The persistent observation of older age in biotype 1 might initially suggest a confounding role. We find this unlikely given the modest age disparities (4-5 years), likely insufficient to explain the observed FNC variations. Analyses for cognitive and FNC differences were age-adjusted, minimizing the likelihood that age alone can account for the results. The disparities in childhood learning difficulties suggest that differences between biotypes are present since childhood and independently of the participants’ current ages. To further validate our results, we conducted k-means clustering again after regressing out the influence of age from FNC features. 92.1% of patients in the discovery and 93.0% in the replication sets were assigned to the same biotype as in the original analyses, additionally supporting that age did not play a major role (supplementary material).

Furthermore, the percentage of relatives in the same cluster, is reduced to 54.85% (χ2=1.22, p=0.269) when using SZ and BPD diagnoses instead of cognitive biotypes, suggesting that SZ and BPD groups are similar or overlapping in CFPs. This suggests that a neurobiologically driven stratification could be a better approach to foster treatment development for cognitive dysfunction, rather than approaches based on DSM diagnoses. Subsequent investigations could explore the impact of treatments purportedly capable of modulating cognition in psychotic disorders (3) on CFPs. This exploration may offer insights into which patients may experience cognitive improvements or deterioration as a pharmacological side effect (43).

### Limitations

First, even though we have a relatively large sample size, it may have been insufficient for some analyses. Second, to avoid simply capturing general population variability in clustering, we selected 1) FNC canonical variates within pairs with differences between participants with psychosis and controls in both canonical variates and 2) FNC features with the highest correlation with this canonical variate in participants with psychosis. However, it still may be possible that identified biotypes correspond to general population variability and not specific to psychotic disorders. Conducting multivariate analyses using only the participants with psychosis may be another approach. Third, hard clustering techniques as k-means present limitations since some participants may present intermediate characteristics between clusters. Fourth, we did not adjust for drug use or medical conditions known to impact cognition (cardiometabolic conditions) (44,45). Fifth, about 60% of our sample were white/caucasian, limiting the generalization of results. Differences in the representation of races in biotypes may not extrapolate outside the US to countries with different social structures.

## CONCLUSIONS

We have identified and, to a reasonable extent, replicated two distinct cognitive biotypes in a large sample of patients with psychotic disorders. These biotypes exhibit disparities in cognitive dysfunction severity, demographics, and brain functional alterations with distinct patterns of hypo-hyperconnectivity. These biotypes may be partially present in first-degree relatives. Utilizing these biotypes as a stratification framework in future investigations focused on cognitive dysfunction may be promising for enhancing their success.

## Data Availability

The storage and management of the data and access procedures are overseen by the National Institute of Mental Health (NIMH) through the National Data Archive (NDA).

https://nda.nih.gov/

