## Supplemental Material for "Neurobiology-based Cognitive Biotypes Using Multi-scale Intrinsic Connectivity Networks in Psychotic Disorders"

**SUPPLEMENTARY METHODS**

**1. Participants**

The storage and management of the data and access procedures are overseen by the National Institute of Mental Health (NIMH) through the National Data Archive (NDA). Detailed instructions for accessing the data have been supplied elsewhere (1). Detailed information on the recruitment strategies can be found elsewhere (1–3). Strategies were consistent for both Bipolar and Schizophrenia Network for Intermediate Phenotypes (B-SNIP) datasets 1 and 2.

The Institutional Review Board approved the project at each participating institution: Athens, GA (University of Georgia, B-SNIP2 only); Baltimore, MD (University of Maryland School of Medicine, B-SNIP1 only); Boston, MA (Harvard Medical School), Chicago IL (University of Illinois-Chicago for B-SNIP1 and University of Chicago for B-SNIP2); Dallas, TX (University of Texas Southwestern Medical Center), Detroit MI (Wayne State University, B-SNIP1 only); and Hartford, CT (Yale University School of Medicine). Before taking part, all participants provided written informed consent after a comprehensive explanation of the study procedures.

2. **Discovery and replication sets**

In the replication set, one of our aims was to examine the influence of covariates, specifically the average daily chlorpromazine dose, on the statistically significant canonical correlations. To ensure the availability of chlorpromazine equivalent data for all patients in the replication set, we randomly selected 40% of patients who have such data available (54.20% of patients in the entire sample have chlorpromazine equivalent data) to be part of the replication set.

3. **Clinical and cognitive assessments**

Demographic data were collected for all participants. Participants underwent the Structured Clinical Interview for Diagnostic and Statistical Manual of Mental Disorders IV. Assessments were conducted when participants were clinically stable. Participants who meet the criteria for a psychotic disorder were rated in Montgomery-Asberg Depression Rating Scale (4), Positive and Negative Syndrome Scale (5), and Young Mania Rating scales (6). All participants were rated on the Hollingshead Two-Factor Socioeconomic Rating Scale (7), Global Assessment of Functioning (8), and Birchwood Social Functioning (9). Missing data in demographic and clinical variables were imputed with chained random forest (<5% of missing data) (10,11). Where possible, the average daily chlorpromazine dose for each subject was computed (45.8% of missing data) (12).

Brief Assessment of Cognition in schizophrenia (BACS) (13,14): It includes six subtests encompassing four cognitive domains: 1) Verbal Memory/List Learning Task, 2) Working Memory/Digit Sequencing Task, 3) Motor Speed/Token Motor Task, 4) Verbal Fluency/Category Instances Task, Controlled Oral Word Association Test, 5) Attention and Speed of processing/Symbol Coding Task, and 6) Executive functions/Tower of London. To gauge overall cognitive impairment in psychotic disorders, a composite score known as the BACS composite score integrates performance across the four domains (14). Normative data stratified by age and sex were employed to compute composite scores for every participant (1,14–16). We checked for outliers or improbable values. Extreme subtest scores were capped at a z-score of -4.0, as previously documented (1,14–16).

4. **Imaging Data Acquisition**

During the scan, participants were instructed to keep their eyes open and focus on a crosshair displayed on a monitor while keeping stillness. All participants at each site underwent a 5-minute rsfMRI on a 3 Tesla MRI scanner. To minimize head motion, a custom-built head-coil cushion was employed. Immediately after the scan, participants' alertness was confirmed, and if necessary, a repeat scan was conducted. These instructions effectively reduced head motion, prevented participants from falling asleep, and served as an experimental control for visual input. These details have been previously reported elsewhere (17).

5. **Imaging Preprocessing**

We employed a combination of FMRIB Software Library (FSL v6.0, <https://fsl.fmrib.ox.ac.uk/fsl/fslwiki/>) and the statistical parametric mapping (SPM12, <http://www.fil.ion.ucl.ac.uk/spm/>) toolboxes within the MATLAB environment to carry out the preprocessing steps. The preprocessing steps include rigid body motion correction, slice timing correction, and distortion correction to address susceptibility-induced distortions resulting from different acquisition parameters. Next, we warped the preprocessed subject data into the Montreal Neurological Institute (MNI) space using an echo-planar imaging (EPI) template (18). Lastly, we resampled the subject data to isotropic voxels of 3mm³ and applied spatial smoothing using a Gaussian kernel with a full width at half-maximum (FWHM) of 6mm.

6. **Extracting subject-specific multi-scale intrinsic connectivity networks (ICNs)**

Multivariate-objective optimization ICA with reference (MOO-ICAR) is a spatially constrained independent component analyses (sc-ICA) approach, which estimates ICNs by optimizing their independence while maintaining their similarity with the reference. We chose MOO-ICAR as it has demonstrated effective performance in capturing subject-specific information while effectively eliminating artifacts (19–21).

7. **Estimating subject-specific functional network connectivity (FNC)**

Prior to calculating subject-specific Functional Network Connectivity (FNC), the ICN time courses underwent an additional post hoc cleaning procedure to mitigate the effect of remaining noise, which may not be wholly removed using ICA (19). The procedure included removing linear, quadratic, and cubic trends, regressing out the six motion realignment parameters and their derivatives, replacing outliers with the best estimate using a third-order spline fit to the clean portions of the time courses, and applying bandpass filtering using a fifth-order Butterworth filter with a cutoff frequency of 0.01 Hz-0.15 Hz. Subsequently, we computed subject-specific FNC by calculating pairwise Pearson correlations between cleaned ICN time courses. This process resulted in a 105 × 105 symmetric FNC matrix. We further applied Fisher's Z transform to improve normality.

Finally, we also regressed out the site-related and head motion effects (mean framewise displacement) in the discovery set. Subsequently, we employed the parameter coefficients derived from the linear model in the discovery set to minimize their influence on the replication set. This approach was chosen to prevent data leakage between the discovery and replication sets.

### 8. **Canonical FNC signatures of cognitive performance**

Canonical correlation analysis (CCA) is a multivariate analysis technique that captures relationships between two sets of data, specifically in our case, the interplay between cognitive performance and FNC features. It finds pairs of weights that create linear combinations that maximize the correlation between the resulting latent variables (canonical variates) (22–26). However, due to a smaller sample size compared to the number of FNC features, standard CCA faces the issue of being ill-posed, leading to non-unique solutions and overfitting the data. Consequently, the obtained results may lack generalizability. To address the high dimensionality problem (5640 FNC features) and multicollinearity (22,23,26,27), Principal Component Analysis (PCA) plus CCA (PCA-CCA) was chosen over standard CCA (22). By transforming data into orthogonal principal components and retaining the top components that explain the most variance, we make our model well-posed and address the potential multicollinearity problem. However, one challenge in this process is potentially losing valuable information for linking the two modalities when discarding principal components with low variance. To tackle this, we estimated the optimal number of principal components directly from the data, ensuring that we retained relevant information while achieving a more generalizable model. To tune the model, find an optimal number of principal components, and improve the robustness of our findings, we randomly split the discovery set into 80% train subset and 20% test subsets, estimated canonical pairs for a given number of principal components using 80% train subset, and estimated the correlation value for canonical pairs in the test subset. We repeated this process 10,000 times for different random splits and selected the number of principal components with the highest average correlation value in the test subsets across splits. We selected the canonical variates with a significant association (p<0.05) in test subsets throughout 10,000 permutations and derived the final canonical variates as a weighted average of these canonical variates, where the weight was the correlation value (ρ) between the canonical variates in each permutation. It is worth noting that the sign of canonical variables is arbitrary; therefore, before calculating the average value, we aligned the signs of canonical variables across permutations.

For linear models, since we were aware of possible problems associated with covariates correlated with one group (28) we repeated these analyses excluding chlorpromazine equivalents (associated with participants with psychosis) and obtained similar results.

### 9. **Identifying cognition-related psychosis biotypes**

We performed k-means clustering with 50 random starts and Euclidean distance (37,38) using patients in the discovery set. Clustering stability was evaluated using a bootstrapping resampling technique (*n* bootstraps=1000). Silhouette index was computed to find the optimal number of clusters (k), and its statistical significance was assessed against the null hypothesis of data coming from a distribution with no underlying clusters (27,29). The code and procedure provided in (27) and employed elsewhere (30,31). K-means clustering was performed on each bootstrapped resample, and Jaccard similarity values were computed (32,33).

**Biotypes validation and characterization**

We assessed the differences in FNC features between biotypes of patients obtained in the discovery set, biotypes of patients obtained by assignment in the replication set, and biotypes of relatives, compared to healthy controls using linear models adjusting for sex, age, race, ethnicity, and head motion and site, and adjusting p-values with False Discovery Rate (significance as p_adjusted_< 0.05).

10. **Correlation between t-statistics for the differences between biotypes in FNC features and loadings for canonical variates**

We wanted to assess if the differences in FNC features between biotypes were larger in FNC features with a larger association with cognition, i.e., if brain characteristics that differentiate our biotypes are related to cognition and therefore biotypes can be considered as different subgroups in brain characteristics linked to cognition. With this purpose, we obtained t-statistics standing for the differences between biotypes in FNC features in the replication set. Next, we computed the correlation between t-statistics and canonical loadings for the first cognitive canonical variate, representing the contribution of each FNC feature to cognitive performance. We repeated the procedure for SZ-BPP between-group differences. We excluded SAD participants to keep the procedure with two groups.

11. **Software**

Analyses were conducted using the R Statistical language (version 4.1.2; R Core Team, 2021), MATLAB Version 2022b, and Python 3.10.10.

Relevant packages:

R: ggplot2, stats, tidymodels, caret, tidyclust, cluster, emmeans, gtsummary, ggstatsplot, NbClust, fpc, MASS.

Python: Network plots were generated using netplotbrain (34).

**SUPPLEMENTARY RESULTS**

### 1. **Canonical FNC signatures of cognitive performance**

In the first canonical pair, age (β=-0.14 CI: -0.21 – -0.06, p<0.001) and age×CV_FNC_ (β=-0.08 CI: -0.14 – -0.01, p=0.027) were statistically significant predictors of CV_Cog_. In the second canonical pair, no interaction was statistically significant. Pairwise comparison in the replication set also showed statistically significant differences in CV_FNC1_ between relatives–patients, d=0.60, t(466)=5.05, p_adj_<0.0001; CV_Cog1_: relatives–patients, d=0.42, t(466)=4.00, p_adj_=0.0002; CV_FNC2_ : controls–relatives, d=-0.36, t(466)=-2.84, p_adj_=0.013; relatives–patients, d=-0.43, t(466)=-3.34, p_adj_=0.003; and CV_Cog2_: relatives–patients, d=0.34, t(466)=2.79, p_adj_=0.02. Assumptions of linear models were adequately met in all models.

2. **Identifying cognition-related psychosis biotypes, validation, and characterization**

367/465 relatives had their family members included in the analyses. 281/367 (76.57%) were assigned to psychosis clusters/biotype; of those, 197/281 (70.12%, χ2=7.0297, p<0.00001) were assigned to the same cluster/biotype as their family member. 249/326 relatives of participants with SZ or BPP had their family members included in the analyses. 175/249 (70.28%) were assigned to BPP or SZ clusters; of those, 96/175 (54.85%, χ2=1.2215, p=0.269) were assigned to the same cluster as their family members.

3. **Correlation between t-statistics for the differences between clusters in FNC features and loadings for canonical variates**

T-statistics of between-biotypes differences in replication set for each FNC feature and after accounting for sex, age, race, and average daily chlorpromazine dose showed a statistically significant Pearson-correlation with CV_FNC1_ canonical loadings (r_1_=0.58, CI_1_: 0.56–0.60, df_1_=5458, p_1_<0.0001), and CV_FNC2_ canonical loadings (r_2_=-0.45, CI_2_: -0.47– -0.42, df_2_=5458, p_2_<0.0001). The same analyses conducted with T-statistics of the differences between patients with Schizophrenia and Bipolar Disorder showed correlations substantially smaller, especially for CV_FNC1_ canonical loadings, the FNC canonical variate with the highest canonical correlation (r_1_=-0.11, CI_1_: -0.14 – -0.09, df_1_=5458, p_1_<0.0001; r_2_=-0.43, CI_2_:-0.46 – -0.41, df_2_=5458, p_2_<0.0001).

4. **Age sensitivity analyses**

After inspecting the results, and due to the consistent difference in age between biotypes, we aimed to analyze the influence of age on clustering. With this purpose, we regressed out the influence of age on FNC in the discovery set and ran k-means clustering again. 92.1% of patients were assigned to the same cluster compared with k-means clustering with no regressing out in age. Additionally, we regressed out the influence of age on FNC in the replication set and repeated the process of assigning patients to one of the clusters obtained from the discovery set with the influence of age regressed out. 93.0% of patients in the replication set were assigned to the same cluster as in the original analyses. We concluded that while age may play a role, its impact on the outcomes of k-means clustering may be minimal.

**SUPPLEMENTARY TABLES**

**Table S1. Demographic, clinical, cognitive, and site characteristics of participants in discovery and replication sets**

|  | | Replication | | | | | | | | | | | | Discovery | | | | | | | | | | |
| --- | --- | --- | --- | --- | --- | --- | --- | --- | --- | --- | --- | --- | --- | --- | --- | --- | --- | --- | --- | --- | --- | --- | --- | --- |
| **Characteristic** | | **Healthy Controls**  **N = 126** | | | | **Relatives**  **N = 93** | | | | **Patients**  **N = 256** | | | | **Healthy Controls**  **N = 500** | | | | **Relatives**  **N = 372** | | | | **Patients**  **N = 923** | | |
| BDP* | |  | | | | 31 | | 33% | | 60 | | 23% | |  | | | | 121 | | 32% | | 250 | | 27% |
| SAD* | |  | | | | 28 | | 30% | | 96 | | 38% | |  | | | | 111 | | 30% | | 299 | | 32% |
| SZ* | |  | | | | 34 | | 37% | | 100 | | 39% | |  | | | | 140 | | 38% | | 374 | | 41% |
| Demographic | | | | | | | | | | | | | | | | | | | | | | | | |
| Age (years) | | 35.55 | | 12.94 | | 41.66 | | 15.88 | | 37.69 | | 11.79 | | 35.92 | | 12.20 | | 41.12 | | 15.59 | | 36.74 | | 12.20 |
| Sex (Female) | | 74 | | 60% | | 63 | | 69% | | 137 | | 54% | | 293 | | 59% | | 237 | | 64% | | 445 | | 48% |
| SES | | 35.70 | | 14.45 | | 38.41 | | 16.24 | | 45.81 | | 14.26 | | 35.54 | | 13.24 | | 39.77 | | 16.66 | | 47.51 | | 14.67 |
| Ethnicity | | | | | | | | | | | | | | | | | | | | | | | | |
| Hispanic | | 14 | | 11% | | 6 | | 6.5% | | 35 | | 14% | | 65 | | 13% | | 39 | | 10% | | 111 | | 12% |
| Not Hispanic | | 112 | | 89% | | 87 | | 94% | | 219 | | 86% | | 435 | | 87% | | 333 | | 90% | | 811 | | 88% |
| Not ascertainable | | 0 | | 0% | | 0 | | 0% | | 2 | | 0.8% | | 0 | | 0% | | 0 | | 0% | | 1 | | 0.1% |
| Race | | | | | | | | | | | | | | | | | | | | | | | | |
| African American | | 33 | | 26% | | 32 | | 34% | | 98 | | 38% | | 167 | | 33% | | 100 | | 27% | | 351 | | 38% |
| Caucasian | | 75 | | 60% | | 61 | | 66% | | 123 | | 48% | | 264 | | 53% | | 255 | | 69% | | 469 | | 51% |
| Other | | 18 | | 14% | | 0 | | 0% | | 35 | | 14% | | 69 | | 14% | | 17 | | 4.6% | | 103 | | 11% |
| Clinical | | | | | | | | | | | | | | | | | | | | | | | | |
| BSFS | | 154.87 | | 15.43 | | 149.20 | | 23.98 | | 122.25 | | 22.76 | | 154.21 | | 17.40 | | 146.51 | | 21.25 | | 124.79 | | 24.61 |
| GAF | | 84.19 | | 6.23 | | 76.72 | | 12.49 | | 54.68 | | 12.47 | | 85.01 | | 6.75 | | 75.67 | | 13.51 | | 53.61 | | 13.30 |
| MADRS | |  | |  | |  | |  | | 12.04 | | 9.72 | |  | |  | |  | |  | | 10.79 | | 9.53 |
| PANSS | |  | |  | |  | |  | | 61.34 | | 18.77 | |  | |  | |  | |  | | 61.14 | | 18.65 |
| SBS | |  | |  | |  | |  | | 5.11 | | 2.62 | |  | |  | |  | |  | | 5.22 | | 2.85 |
| YMRS | |  | |  | |  | |  | | 8.31 | | 6.67 | |  | |  | |  | |  | | 8.04 | | 7.45 |
| Cognition | | | | | | | | | | | | | | | | | | | | | | | | |
| BACS Composite Score | | -0.17 | | 1.18 | | -0.54 | | 1.25 | | -1.48 | | 1.39 | | -0.26 | | 1.21 | | -0.48 | | 1.32 | | -1.50 | | 1.39 |
| BACS Verbal Memory | | -0.11 | | 1.13 | | -0.27 | | 1.26 | | -1.01 | | 1.33 | | -0.21 | | 1.20 | | -0.36 | | 1.22 | | -1.02 | | 1.40 |
| BACS Digit Sequencing | | -0.24 | | 1.03 | | -0.31 | | 1.18 | | -0.89 | | 1.17 | | -0.22 | | 1.06 | | -0.38 | | 1.24 | | -0.96 | | 1.24 |
| BACS Token Motor | | -0.33 | | 1.11 | | -0.69 | | 0.99 | | -1.52 | | 1.28 | | -0.49 | | 1.22 | | -0.36 | | 1.16 | | -1.44 | | 1.16 |
| BACS Verbal Fluency | | 0.14 | | 1.13 | | -0.11 | | 1.29 | | -0.53 | | 1.18 | | 0.14 | | 1.12 | | -0.11 | | 1.12 | | -0.46 | | 1.20 |
| BACS Symbol Coding | | -0.08 | | 1.00 | | -0.49 | | 1.09 | | -1.08 | | 1.20 | | -0.14 | | 1.03 | | -0.46 | | 1.08 | | -1.16 | | 1.12 |
| BACS Tower of London | | 0.12 | | 0.90 | | -0.14 | | 0.96 | | -0.41 | | 1.09 | | 0.01 | | 0.96 | | -0.11 | | 1.00 | | -0.41 | | 1.04 |
| WMS Spatial Span Forward | | 8.40 | | 2.08 | | 7.96 | | 2.06 | | 7.52 | | 2.07 | | 8.42 | | 2.05 | | 7.88 | | 2.03 | | 7.48 | | 2.09 |
| WMS Spatial Span Backward | | 7.89 | | 2.08 | | 6.82 | | 2.18 | | 6.53 | | 2.14 | | 7.68 | | 2.02 | | 7.05 | | 2.15 | | 6.47 | | 2.23 |
| Medication | | | | | | | | | | | | | | | | | | | | | | | | |
| ChlorEq | |  | | | |  | | | | 357.40 | | 350.16 | |  | | | |  | | | | 365.34 | | 346.73 |
| Missing | |  | | | |  | | | | 0 | | | |  | | | |  | | | | 540 | | |
| Site | | | | | | | | | | | | | | | | | | | | | | | | |
| Baltimore | 9 | | 7.1% | | 21 | | 23% | | 18 | | 7.0% | | 42 | | 8.4% | | 87 | | 23% | | 105 | | 11% | |
| Boston | 8 | | 6.3% | | 2 | | 2.2% | | 15 | | 5.9% | | 26 | | 5.2% | | 9 | | 2.4% | | 68 | | 7.4% | |
| Chicago | 32 | | 25% | | 12 | | 13% | | 58 | | 23% | | 113 | | 23% | | 64 | | 17% | | 229 | | 25% | |
| Dallas | 18 | | 14% | | 13 | | 14% | | 47 | | 18% | | 96 | | 19% | | 59 | | 16% | | 145 | | 16% | |
| Detroit | 7 | | 5.6% | | 7 | | 7.5% | | 15 | | 5.9% | | 15 | | 3.0% | | 30 | | 8.1% | | 36 | | 3.9% | |
| Georgia | 22 | | 17% | | 0 | | 0% | | 26 | | 10% | | 81 | | 16% | | 0 | | 0% | | 89 | | 9.6% | |
| Hartford | 30 | | 24% | | 38 | | 41% | | 77 | | 30% | | 127 | | 25% | | 123 | | 33% | | 251 | | 27% | |
| Dataset | | | | | | | | | | | | | | | | | | | | | | | | |
| B-SNIP 1 | 47 | | 37% | | 93 | | 100% | | 95 | | 37% | | 188 | | 38% | | 372 | | 100% | | 399 | | 43% | |
| B-SNIP 2 | 79 | | 63% | | 0 | | 0% | | 151 | | 63% | | 312 | | 62% | | 0 | | 0% | | 524 | | 57% | |

**Table S1:** * For relatives it is the diagnosis of their affected family member; SES: Socioeconomic Status; MADRS: Montgomery-Asberg Depression Rating Scale; YMRS: Young Mania Rating Scale; PANSS: Positive and Negative Syndrome Scale for Schizophrenia; BSFS: Birchwood Social Functioning Scale; GAF: Global Assessment of Functioning Scale; SBS: Schizo-bipolar Scale; BACS: Brief Assessment of Cognition in Schizophrenia; WMS Weschler Memory Scale; ChlorEq: average daily chlorpromazine dose

**Table S2. Linear models for each canonical pair in the replication set**

|  | **CV_Cog1_** | | | **CV_Cog2_** | | | **CV_Cog3_** | | |
| --- | --- | --- | --- | --- | --- | --- | --- | --- | --- |
| Predictors | β | CI | p | β | CI | p | β | CI | p |
| (Intercept) | -0.06 | -0.24 – 0.12 | 0.496 | 0.20 | -0.04 – 0.43 | 0.101 | 0.40 | 0.13 – 0.66 | **0.004** |
| CV_FNC1_ | 0.24 | 0.16 – 0.33 | **<0.001** |  |  |  |  |  |  |
| Age | -0.14 | -0.21 – -0.06 | **<0.001** | 0.26 | 0.17 – 0.35 | **<0.001** | 0.12 | 0.02 – 0.21 | **0.018** |
| Sex (male) | 0.12 | -0.03 – 0.27 | 0.114 | 0.13 | -0.04 – 0.30 | 0.140 | -0.36 | -0.54 – -0.18 | **<0.001** |
| Race (Other) | 0.14 | -0.14 – 0.41 | 0.314 | 0.05 | -0.24 – 0.33 | 0.743 | -0.35 | -0.68 – -0.03 | **0.033** |
| Race (Caucasian) | 0.44 | 0.28 – 0.60 | **<0.001** | -0.09 | -0.28 – 0.10 | 0.348 | -0.31 | -0.52 – -0.10 | **0.003** |
| SES | -0.25 | -0.33 – -0.17 | **<0.001** | 0.13 | 0.04 – 0.21 | **0.004** | -0.03 | -0.13 – 0.06 | 0.489 |
| ChlorEq | -0.12 | -0.22 – -0.04 | **0.008** | -0.10 | -0.21 – 0.02 | 0.093 | -0.02 | -0.13 – 0.08 | 0.740 |
| Group (relatives) | -0.23 | -0.42 – -0.03 | **0.020** | -0.06 | -0.31 – 0.20 | 0.663 | -0.01 | -0.28 – 0.27 | 0.962 |
| Group (patients) | -0.39 | -0.59 – -0.19 | **0.001** | -0.36 | -0.61 – -0.12 | **0.005** | -0.07 | -0.33 – 0.18 | 0.561 |
| CV_FNC1_× Age | -0.08 | -0.14 – -0.01 | **0.027** |  |  |  |  |  |  |
| CV_FNC2_ |  |  |  | 0.12 | 0.02 – 0.21 | **0.015** |  |  |  |
| CV_FNC3_ |  |  |  |  |  |  | 0.03 | -0.07 – 0.14 | 0.500 |
| Observations | 475 | | | 475 | | | 475 | | |
| R^2^ / R^2^ adjusted | 0.419 / 0.406 | | | 0.157 / 0.141 | | | 0.091 / 0.073 | | |

**Table S2:** CV_cog_: Cognitive canonical variate. CV_FNC_: Functional network connectivity canonical variate. SES: Socioeconomic status. ChlorEq: Average daily chlorpromazine dose. CI: Confident interval. Only significant interactions were kept in the final models.

**Table S3. Comparison of demographic, clinical, and cognitive characteristics of biotypes**

| Patients’ discovery set | | | | | | | | | |
| --- | --- | --- | --- | --- | --- | --- | --- | --- | --- |
|  | Biotype 1  N=426 | | Biotype 2  N=497 | | Analyses | | | | |
|  | Estimated Marginal Mean | SE | Estimated Marginal Mean | SE | d | t | df | p | q |
| Age (years) | 38.90 | 0.58 | 34.90 | 0.54 | 3.92 | 4.931 | 921 | **<0.0001** | **<0.001** |
| ChlorEq | 492 | 48.70 | 387 | 46.30 | 105 | 1.564 | 381 | 0.119 | 0.147 |
| BFSF | 121 | 1.27 | 128 | 1.16 | -6.10 | -3.553 | 813 | **<0.0001** | **<0.001** |
| GAF | 52.10 | 0.64 | 54.90 | 0.60 | -2.75 | -3.145 | 921 | **0.002** | **0.004** |
| MADRS | 10.90 | 0.46 | 10.70 | 0.43 | 0.121 | 0.193 | 921 | 0.847 | 0.847 |
| PANSS | 61.30 | 0.90 | 61.0 | 0.84 | 0.29 | 0.235 | 921 | 0.813 | 0.847 |
| SBS | 5.64 | 0.14 | 4.85 | 0.13 | 0.792 | 4.240 | 921 | **<0.0001** | **<0.001** |
| SES | 50.0 | 0.70 | 45.40 | 0.65 | 4.60 | 4.802 | 921 | **<0.0001** | **<0.001** |
| YMRS | 7.82 | 0.36 | 8.22 | 0.33 | -0.41 | -0.830 | 921 | 0.406 | 0.467 |
| BACS Composite Score^b^ | -1.76 | 0.44 | -1.14 | 0.43 | -0.61 | -6.568 | 911 | **<0.0001** | **<0.001** |
| BACS Verbal Memory^b^ | -1.89 | 0.45 | -1.38 | 0.43 | -0.46 | -5.338 | 911 | **<0.0001** | **<0.001** |
| BACS Digit Sequencing^b^ | -1.49 | 0.4 | -1.06 | 0.4 | -0.4 | -5.01 | 911 | **<0.0001** | **<0.001** |
| BACS Token Motor^b^ | -0.79 | 0.39 | -0.52 | 0.39 | -0.27 | -3.219 | 905 | **0.0013** | **0.003** |
| BACS Verbal Fluency^b^ | -0.57 | 0.39 | -0.26 | 0.39 | -0.32 | -3.764 | 910 | **0.0002** | **<0.001** |
| BACS Symbol Coding^b^ | -1.12 | 0.36 | -0.67 | 0.36 | -0.46 | -5.839 | 911 | **<0.0001** | **<0.001** |
| BACS Tower of London^b^ | -0.81 | 0.34 | -0.50 | 0.33 | -0.30 | -4.17 | 906 | **<0.0001** | **<0.001** |
| WMS Spatial Span Backward^a^ | -0.16 | 0.29 | 0.14 | 0.28 | -0.29 | -4.587 | 901 | **<0.0001** | **<0.001** |
| WMS Spatial Span Forward^a^ | 0.03 | 0.32 | 0.25 | 0.31 | -0.22 | -3.190 | 902 | **0.0015** | **0.003** |
|  | N | % | N | % | Difference | χ^2^ | df | p | q |
| Special Education | 65/126 | 34% | 67/238 | 22% | 12% | 8.146 | 1 | **0.004** | **0.007** |
| Summer School | 79 | 42% | 92 | 30% | 12% | 6.806 | 1 | **0.009** | **0.013** |
| Repeated Grade | 54 | 29% | 60 | 20% | 9% | 4.925 | 1 | **0.026** | **0.036** |
| Difficulties Learning Maths | 56 | 30% | 57 | 19% | 11% | 8.343 | 1 | **0.004** | **0.007** |
| Difficulties Learning to Read | 88 | 48% | 104 | 34% | 14% | 7.762 | 1 | **0.005** | **0.008** |
| African American | 209 | 49% | 142 | 29% | 20% | 40.000 | 1 | **<0.0001** | **<0.001** |
| Caucasian | 174 | 41% | 295 | 59% | -18% | 30.712 | 1 | **<0.0001** | **<0.001** |
| Other races | 43 | 10% | 60 | 12% | -2% | 0.717 | 1 | 0.397 | 0.467 |
| Hispanic | 47 | 11% | 64 | 13% | -2% | 0.574 | 1 | 0.449 | 0.497 |
| Sex (female) | 219 | 51% | 227 | 46% | 5% | 2.795 | 1 | 0.094 | 0.122 |
| DSM Diagnosis |  |  |  |  |  |  |  |  |  |
| Schizophrenia | 193 | 45% | 181 | 36% | 9% | 7.150 | 1 | **0.008** | **0.012** |
| Schizoaffective Disorder | 140 | 33% | 159 | 32% | 1% | 0.050 | 1 | 0.832 | 0.847 |
| Bipolar disorder | 93 | 22% | 157 | 32% | -10% | 10.570 | 1 | **0.001** | **0.002** |
| Patients’ replication set \| \| \| \| \| \| \| \| \|; \| Cluster 1 N = 110 \| \| Cluster 2 N = 146 \| \| Analyses \| \| \| \|; \| Estimated Marginal Mean \| SE \| Estimated Marginal Mean \| SE \| d \| t \| df \| p \| q; Age (years) \| 40.40 \| 1.10 \| 35.70 \| 0.96 \| 4.68 \| 3.198 \| 254 \| **0.002** \| 0.062; ChlorpEq \| 622 \| 96.30 \| 430 \| 83.60 \| 192 \| 1.508 \| 254 \| 0.133 \| 0.317; BFSF \| 122 \| 2.32 \| 122 \| 1.95 \| 0.344 \| 0.114 \| 232 \| 0.909 \| 0.939; GAF \| 54.10 \| 1.19 \| 55.10 \| 1.03 \| -0.97 \| -0.618 \| 254 \| 0.537 \| 0.684; MADRS \| 11.90 \| 0.93 \| 12.10 \| 0.81 \| -0.19 \| -0.151 \| 254 \| 0.880 \| 0.939; PANSS \| 62.10 \| 1.79 \| 60.80 \| 1.56 \| 1.25 \| 0.528 \| 254 \| 0.598 \| 0.713; SBS \| 5.45 \| 0.25 \| 4.86 \| 0.22 \| 0.60 \| 1.815 \| 254 \| 0.070 \| 0.271; SES \| 47.20 \| 1.36 \| 44.80 \| 1.18 \| 2.42 \| 1.344 \| 254 \| 0.180 \| 0.351; YMRS \| 8.15 \| 0.64 \| 8.43 \| 0.55 \| -0.29 \| -0.339 \| 254 \| 0.735 \| 0.813; BACS Composite Score^b^ \| -1.58 \| 0.33 \| -1.22 \| 0.33 \| -0.35 \| -2.104 \| 244 \| **0.036** \| 0.186; BACS Verbal Memory^b^ \| -0.73 \| 0.32 \| -0.61 \| 0.32 \| -0.13 \| -0.779 \| 244 \| 0.436 \| 0.675; BACS Digit Sequencing^b^ \| -0.97 \| 0.29 \| -0.74 \| 0.29 \| -0.23 \| -1.574 \| 244 \| 0.117 \| 0.302; BACS Token Motor^b^ \| -1.74 \| 0.33 \| -1.68 \| 0.33 \| -0.06 \| -0.370 \| 242 \| 0.712 \| 0.81; BACS Verbal Fluency^b^ \| -0.50 \| 0.30 \| -0.25 \| 0.30 \| -0.25 \| -1.624 \| 243 \| 0.105 \| 0.295; BACS Symbol Coding^b^ \| -1.26 \| 0.30 \| -0.97 \| 0.3 \| -0.29 \| -1.935 \| 243 \| 0.054 \| 0.239; BACS Tower of London^b^ \| -0.45 \| 0.26 \| -0.07 \| 0.26 \| -0.38 \| -2.872 \| 243 \| **0.004** \| 0.062; WMS Spatial Span Backward^a^ \| -0.17 \| 0.21 \| 0.12 \| 0.22 \| -0.29 \| -2.625 \| 238 \| **0.009** \| 0.093; WMS Spatial Span Forward^a^ \| -0.06 \| 0.23 \| 0.21 \| 0.23 \| -0.28 \| -2.364 \| 238 \| **0.018** \| 0.112; \| N \| % \| N \| % \| Difference \| χ^2^ \| df \| p \| q; Special Education \| 20 \| 33% \| 23 \| 26% \| 11% \| 0.649 \| 1 \| 0.420 \| 0.643; Summer School \| 24 \| 40% \| 23 \| 26% \| 14% \| 2.703 \| 1 \| 0.100 \| 0.296; Repeated Grade \| 18 \| 30% \| 21 \| 24% \| 6% \| 0.412 \| 1 \| 0.521 \| 0.658; Difficulties Learning Maths \| 22 \| 37% \| 23 \| 26% \| 11% \| 0.354 \| 1 \| 0.552 \| 0.658; Difficulties Learning to Read \| 26 \| 43% \| 33 \| 37% \| 65 \| 1.511 \| 1 \| 0.219 \| 0.377; African American \| 52 \| 47% \| 46 \| 32% \| 15% \| 5.949 \| 1 \| **0.015** \| 0.114; Caucasian \| 47 \| 43% \| 76 \| 52% \| -9% \| 1.828 \| 1 \| 0.176 \| 0.352; Other races \| 11 \| 10% \| 24 \| 16% \| -6% \| 1.691 \| 1 \| 0.193 \| 0.352; Hispanic \| 11 \| 10% \| 24 \| 16% \| -6% \| 1.692 \| 1 \| 0.193 \| 0.352; Sex (female) \| 56 \| 51% \| 82 \| 56% \| -5% \| 0.502 \| 1 \| 0.479 \| 0.658; DSM Diagnosis \| \| \| \| \| \| \| \| \|; Schizophrenia \| 50 \| 45% \| 37 \| 25% \| 20% \| 2.857 \| 1 \| 0.091 \| 0.300; Schizoaffective Disorder \| 37 \| 34% \| 59 \| 40% \| -6% \| 0.956 \| 1 \| 0.328 \| 0.535; Bipolar disorder \| 23 \| 21% \| 50 \| 34% \| -13% \| 0.462 \| 1 \| 0.497 \| 0.658; (Relatives \| \| \| \| \| \| \| \| \| \|; \| Biotype 1 N=153 \| \| Biotype 2 N=178 \| \| Biotype Healthy N=95 \| \| Analyses \| \| \|; \| Estimated Marginal Mean \| SE \| Estimated Marginal Mean \| SE \| Estimated Marginal Mean \| SE \| F \| df \| p \| q; Age (years) \| 44.29 \| 1.26 \| 39.4 \| 1.16 \| 40.8 \| 1.59 \| 5.368 \| 462 \| **0.005^c^** \| **0.018**; BFSF \| 146 \| 1.90 \| 150 \| 1.83 \| 152 \| 2.42 \| 2.318 \| 356 \| 0.100 \| 0.210; GAF \| 75.5 \| 0.96 \| 80.0 \| 0.89 \| 77.3 \| 1.21 \| 6.12 \| 462 \| **0.0024^d^** \| **0.016**; SES \| 39.7 \| 1.33 \| 38.1 \| 1.23 \| 37.0 \| 1.68 \| 0.88 \| 462 \| 0.416 \| 0.513; BACS Composite Score^b^ \| -1.29 \| 0.15 \| -0.65 \| 0.15 \| -0.67 \| 0.16 \| 7.29 \| 454 \| **0.0008^e^** \| **0.008**; BACS Verbal Memory^b^ \| -0.80 \| 0.15 \| -0.44 \| 0.15 \| -0.373 \| 0.16 \| 5.012 \| 453 \| **0.007^f^** \| **0.018**; BACS Digit Sequencing^b^ \| -1.06 \| 0.14 \| -0.733 \| 0.145 \| -0.61 \| 0.16 \| 5.34 \| 450 \| **0.005^g^** \| **0.018**; BACS Token Motor^b^ \| -0.55 \| 0.15 \| -0.36 \| 0.15 \| -0.50 \| 0.16 \| 0.933 \| 452 \| 0.394 \| 0.513; BACS Verbal Fluency^b^ \| -0.55 \| 0.15 \| -0.39 \| 0.15 \| -0.39 \| 0.16 \| 0.82 \| 452 \| 0.440 \| 0.510; BACS Symbol Coding^b^ \| -0.69 \| 0.13 \| -0.30 \| 0.13 \| -0.37 \| 0.14 \| 5.400 \| 454 \| **0.0048^h^** \| **0.018**; BACS Tower of London^b^ \| -0.49 \| 0.12 \| -0.28 \| 0.12 \| -0.32 \| 0.13 \| 1.927 \| 453 \| 0.147 \| 0.281; WMS Spatial Span Backward^a^ \| -0.22 \| 0.11 \| 0.10 \| 0.11 \| -0.013 \| 0.12 \| 5.14 \| 447 \| **<0.006^i^** \| **0.018**; WMS Spatial Span Forward^a^ \| -0.27 \| 0.12 \| 0.28 \| 0.12 \| 0.05 \| 0.13 \| 11.32 \| 447 \| **<0.0001^j^** \| **0.002**; \| N \| % \| N \| % \| N \| % \| χ^2^ \| df \| p \| q; African American \| 50 \| 33% \| 26 \| 15% \| 33 \| 35% \| 8.771 \| 2 \| **0.012^k^** \| **0.030**; Caucasian \| 98 \| 64% \| 142 \| 80% \| 60 \| 63% \| 2.637 \| 2 \| 0.267 \| 0.467; Other races \| 5 \| 3.3% \| 10 \| 5.6% \| 2 \| 2.1% \| 1.725 \| 2 \| 0.422 \| 0.513; Hispanic \| 14 \| 9.2% \| 15 \| 8.4% \| 9 \| 9.5% \| 0.072 \| 2 \| 0.964 \| 0.964; Sex (female) \| 96 \| 63% \| 128 \| 72% \| 61 \| 64% \| 0.733 \| 2 \| 0.692 \| 0.764; DSM Diagnosis \| \| \| \| \| \| \| \| \| \|; Schizophrenia \| 64 \| 42% \| 56 \| 31% \| 32 \| 34% \| 1.813 \| 2 \| 0.404 \| 0.513; Schizoaffective Disorder \| 44 \| 29% \| 56 \| 31% \| 25 \| 26% \| 0.441 \| 2 \| 0.801 \| 0.841; Bipolar disorder \| 45 \| 29% \| 66 \| 37% \| 38 \| 40% \| 1.830 \| 2 \| 0.405 \| 0.51) \| \| \| \| \| \| \| \| \| | | | | | | | | | |

**Table S3:** SES: Socioeconomic status; MADRS: Montgomery-Asberg Depression Rating Scale; YMRS: Young Mania Rating Scale; PANSS: Positive and Negative Syndrome Scale for Schizophrenia; BSFS: Birchwood Social Functioning Scale; GAF: Global Assessment of Functioning Scale; SBS: Schizo-bipolar Scale; BACS: Brief Assessment of Cognition in Schizophrenia; WMS: Weschler Memory Scale; ChlorpEq: Average daily chlorpromazine dose; SE: Standard error; t: Two-tailed t-statistic; d: Difference; df; Degrees of freedom; χ2: Pearson’s Chi-squared statistic; p: Non-adjusted p-value; q: False discovery rate correction for multiple testing p-value. ^a^Statistics were obtained from linear models that also accounted for the influence of age, sex, race, ethnicity, and site. ^b^Statistics were obtained from linear models that also accounted for the influence of race, ethnicity, and site (age and sex accounted for when computing z-scores). ^c^Tukey method for comparing a family of three estimates (p) and FDR for multiple testing (q): Cluster 1 > Cluster 2, t=3.220, p=0.004; q=0.009. ^d^Tukey method for comparing a family of three estimates (p) and FDR for multiple testing (q): Cluster 1 < Cluster 2, t=-3.467, p=0.0017, q=0.006. ^e^Tukey method for comparing a family of three estimates (p) and FDR for multiple testing (q): Cluster 1 < Cluster 2, t=-0.345, p=0.002; q=0.006; Cluster 1 < Cluster HC, t=-3.097, p=0.006, q=0.009. ^f^Tukey method for comparing a family of three estimates (p) and FDR for multiple testing (q): Cluster 1 < Cluster 2, t=-2.546, p = 0.030, q= 0.035; Cluster 1 < Cluster HC, t=-2.879, p=0.012, q=0.016. ^g^Tukey method for comparing a family of three estimates (p) and FDR for multiple testing (q): Cluster 1 < Cluster 2, t=-2.362, p=0.049, q=0.049; Cluster 1 < Cluster HC, t=-3.119, p=0.006, q=0.009. ^h^Tukey method for comparing a family of three estimates (p) and FDR for multiple testing (q): Cluster 1 < Cluster 2, t=-3.104, p=0.006, q=0.009: Cluster 1 < Cluster HC, t =-2.44, p=0.039, q=0.042. ^i^Tukey method for comparing a family of three estimates (p) and FDR for multiple testing (q): Cluster 1 < Cluster 2, t=-3.120, p=0.006, q=0.009. ^j^Tukey method for comparing a family of three estimates (p) and FDR for multiple testing (q): Cluster 1 < Cluster 2, t=-4.736, p<0.0001, q=0.001, Cluster 1 < Cluster HC, t=-2.71, p=0.019, q=0.024. ^k^FDR for comparing three groups (p) and FDR for multiple testing (q): Cluster 1 > Cluster 2, χ2= 14.189, p=0.0001, q=0.001; Cluster 2 < Cluster HC, χ2= 13.652, p=0.0001, q=0.001.
